# The Structural Genome Shapes the Chronic Wound Microbiome

**DOI:** 10.64898/2026.10.13.26365461

**Authors:** Khalid Omeir, Jacob Ancira, Rebecca Gabrilska, Craig Tipton, Nicole Phillips, Joseph Wolcott, Caleb D. Phillips

## Abstract

Recent studies report associations between the patient genome and the chronic wound microbiome, suggesting a causal relationship. Guided by the expectation that larger genomic perturbations are less likely to be neutral, here we study the association of genomic copy number variants (CNVs) with abundances of wound species. Genomic and metagenomic data were collected from 597 patients with chronic wounds at initial clinical visit. Association studies included 831 CNVs and 68 species, implementing a two-cohort design and study-wide significance threshold. There were 47 CNVs (5.7%) associated with 25 species (36.8%); an additional 1,730 associations achieved study-wide significance in one cohort. Associations with haplotype dosage variation dominated significant associations (69.4%) indicating haplotype dosage is important beyond simply locus copy count. Significant co-occurrences between associated and non-associated species, all involving anaerobes, support a model in which the genome selects for certain species that in turn shape the broader microbiome through ecological interactions. Significant CNVs encompassed 66 genes which were functionally overrepresented for several terms, with extracellular matrix (ECM) and ECM-cell interaction terms emphasized. Four collagen and collagen remodeling genes were distributed across four unlinked significant CNVs, a result that strikingly aligns with the previous studies reporting other collagen or ECM component associations. Related, cytoskeletal dynamics are also strongly implicated. Heritability estimates drawing on all available data and controlling for species correlation and patient factors yielded 30 species with significant heritabilities, 24 of which were greater than 20%. These findings extend the genomic basis of the chronic wound microbiome by emphasizing importance of genomic structural variation and suggest a prominent role for variation across ECM-cellular connections as mechanisms defining microbial selection.

## Introduction

Chronic wounds are identified as wounds exhibiting delayed healing, frequently persisting over several months. Whereas there is no universally accepted time-based definition of chronicity^1,2^, chronic wounds, including diabetic foot ulcers, venous leg ulcers, decubitus ulcers, and non-healing surgical wounds, have 12-week healing rates that often do not exceed 40%^3,4^. Despite adherence to current recommended best practices^5^, many chronic wounds fail to heal altogether^6^. The prevalence of chronic wounds is increasing; in the United States, chronic wounds affect approximately 10.5 million Medicare beneficiaries^7^. National wound care expenditures across the United States have been estimated at $150 billion annually^8^. Patients with chronic wounds often present with high rates of morbidity, including diabetes, obesity, and renal disease, which further impair healing^9,10^. Chronic wounds additionally elevate the risk of amputations, which are associated with increased mortality^11,12^. Thus, understanding and addressing the chronic wound epidemic is fundamentally important to societal and individual welfare.

The chronic wound microbiome is now understood to be an important contributor to wound healing. The chronic wound environment is nutrient-rich and conducive to microbial growth. Because microbial contamination pervades chronic wounds, bacterial colonization commonly progresses to infection, causing further tissue damage and exacerbating chronicity^13^. Managing wound bioburden and high-risk microbial taxa is important for improving outcomes and is recommended in wound care guidelines^14–16^. Whereas comprehensively characterizing wound microbial communities is not possible with routine culture, nucleic acid sequencing can and is agnostic to cultivation and aerotolerance^17,18^. Further, guided antimicrobial treatment is associated with better outcomes compared to empirical treatment^19,20^.

Wound microbiomes vary widely in composition across patients^21^. However, the magnitude and relationships among environmental and patient factors contributing to this variability remain poorly understood. Recent work supports that individual genetics influences both microbiome composition and healing. A few two-cohort genomic^22,23^ and transcriptomic^24^ association studies have collectively identified more than 300 human genomic loci associated with either microbial composition^22^ or a diverse set of 48 species^23,24^. Genes at these loci were enriched for pathways relevant to the wound environment and microbial interactions, including extracellular matrix organization (ECM) and signal transduction. In important related work drawing from the same population, differences in wound microbiome composition were strongly predictive of healing outcomes across patients^25^. Thus, the overarching current working hypothesis is that patient-specific genetics shape the wound microenvironment, which in turn influences microbiome composition and ultimately healing trajectories.

Among forms of host genetic variation, copy number variants (CNVs) are deletions or duplications that can be inherited or arise de novo^26^. CNVs are an important class of genomic variants that by spanning large genomic intervals can physically disrupt genes, alter the dosage of genes or regulatory elements^27,28^, making them less likely to be neutral as compared to point mutations. CNVs are thought to commonly arise by nonallelic homologous recombination between duplicated segments^29^, a process that can occur when duplicated segments line up incorrectly during meiosis and recombine to generate a deletion on one chromosome and a reciprocal duplication on the other^30^. Large population analyses of the UK Biobank report CNVs associating with a broad range of human traits and diseases^31^. CNVs are common across the human genome, and high-throughput CNV detection has revealed many CNVs of unknown clinical significance^30^. Previous studies support that CNVs influence microbial variation in the oral cavity^32^, gut^33^, nasopharynx^34^, and vagina^35^, but their contribution to the chronic wound microbiome remains unknown.

Here, using a haplotype-aware framework, we tested CNV dosage and haplotype-set associations with chronic wound microbial abundances in a two-cohort discovery and replication design. The primary objective was to determine whether structural genomic variation contributes to interpatient differences in the chronic wound microbiome and identify the host biological processes implicated by these associations. We demonstrate that structural genomic variation reliably associates with wound species abundances, and these regions repeatedly implicate ECM-related functions. This approach extends investigation of host genomic influences on the chronic wound microbiome, expanding the emerging perspective on genome-microbiome interaction. The longer-term motivation is that defining host genetic factors that predispose patients to microbes and healing will enable prediction useful to guide intervention and treatment.

## Results

### Clinical characteristics and wound microbiome composition

The initial dataset consisted of 597 chronic wound patients. After excluding patients who failed quality control or data missingness thresholds, 482 patients remained (Fig 1A). Patients were predominantly older, male, obese, diabetic, self-reported as White, and nonsmoking (Table 1). 68 bacterial species were present in at least 5 patients (1% prevalence). Across these species, prevalence ranged from 1.04% to 33.82%, with a median of 2.59%. Species were classified by aerotolerance as anaerobic (28), facultative (35), or aerobic (5), and by morphology as Gram-positive (39) or Gram-negative (29; S1 Table).

**Fig 1.**
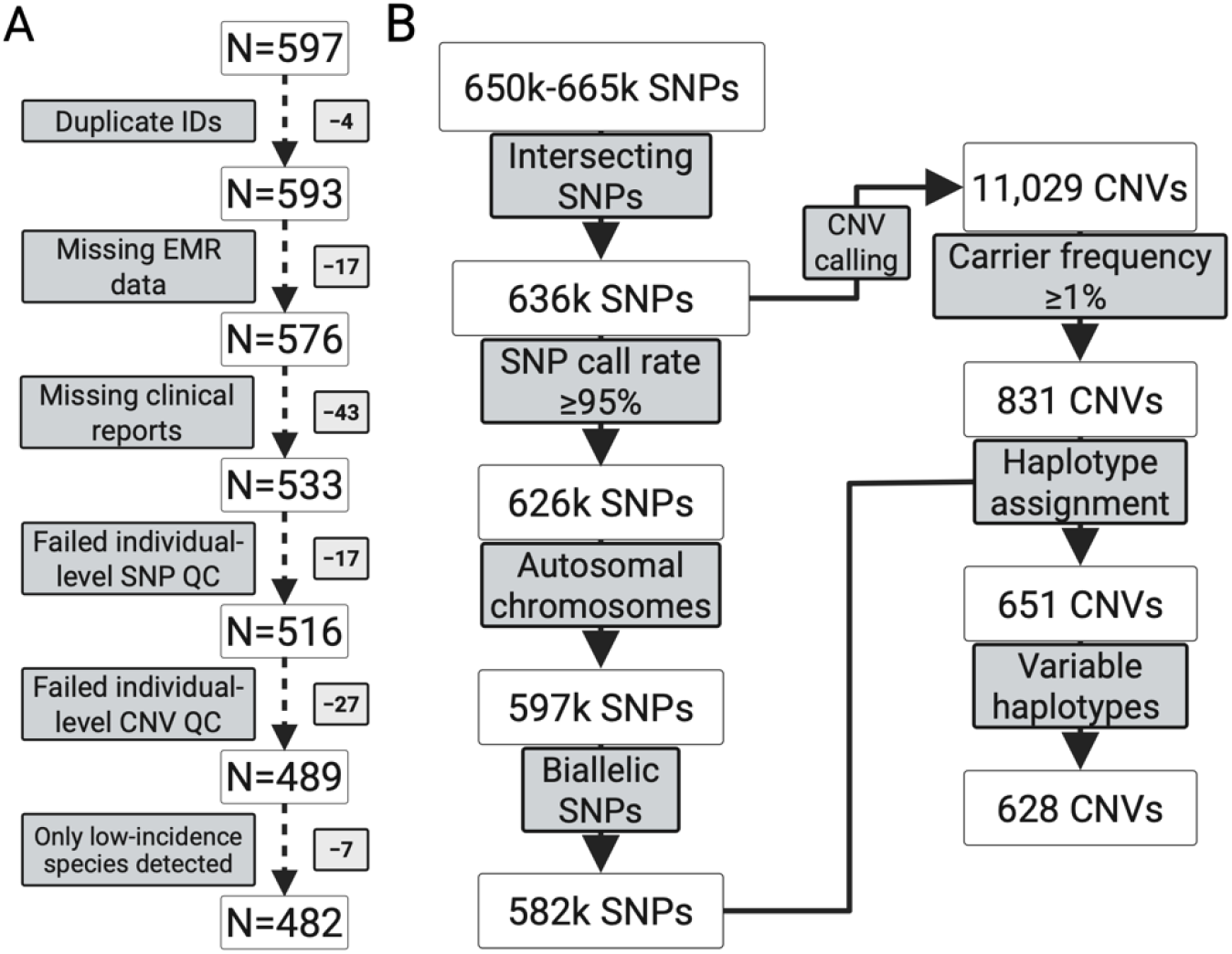
Flowcharts summarizing individual-level quality control and genomic data processing. (A) Sample-level exclusions resulted in a final cohort size of 482 individuals. Boxes on the right indicate the number of individuals excluded at each step. (B) Genotype data from array versions were merged using intersecting SNPs, resulting in 636k shared SNPs used for CNV calling. CNV calls required support from at least three SNP markers. Subsequent SNP-level filtering for haplotype assignment retained 582k SNPs. Of 831 retained CNVs, 651 had sufficient information for haplotype assignment, of which 628 were variable. Created with BioRender.com

**Table 1:** Summary of patient characteristics.

| Characteristic |  |
| --- | --- |
| Total sample size | 482 |
| Age | 66.7 ± 13.2 years |
| Weight | 214.4 ± 63.6 lbs |
| Height | 67.8 ± 5.5 in |
| Self-reported race/ethnicity | White: 308 (63.9%)<br>Hispanic: 139 (28.8%)<br>Black: 35 (7.3%) |
| Sex | Male: 292 (60.6%)<br>Female: 190 (39.4%) |
| Smoker | 89 (18.5%) |
| Diabetic | 307 (63.7%) |
| BMI category | Obese: 252 (52.3%)<br>Overweight: 145 (30.1%)<br>Normal: 70 (14.5%)<br>Underweight: 11 (2.3%)<br>Missing: 4 (0.8%) |

### Genomic data and CNV calling

Marker counts initially ranged from 650,181 to 665,608 across array versions. A total of 635,996 single nucleotide polymorphisms (SNPs) were retained as intersecting markers across array versions, with an overall call rate of 0.99. After SNP-level quality control, 581,697 SNPs were available for phasing and haplotype assignment. Across all samples, 11,029 distinct CNVs were identified, of which 831 were present in at least 1% of the cohort and retained for association testing. Among these, 180 lacked informative SNPs and 23 lacked haplotype variation, so these 203 CNVs were retained for CNV dosage testing only. The remaining 628 CNVs were eligible for haplotype-set association testing (Fig 1B; S2 Table).

### CNV landscape

In our population, CNV count was highest on chromosome 19, followed by chromosome 22 and chromosome 1, and after adjusting for chromosome length, chromosomes 19, 22, and 16 showed the highest CNV densities at 17.3, 15.0, and 6.3 CNVs per 10 Mb, respectively (Fig 2A). CNV density varied significantly after controlling for chromosomal length (Poisson likelihood-ratio test, *p* < 2.2 x 10^-16^). CNVs retained for association testing had a median length of 3.1 kb, whereas those with successful haplotype assignment were longer overall, with a median length of 6 kb (Fig 2B).

**Fig 2.**
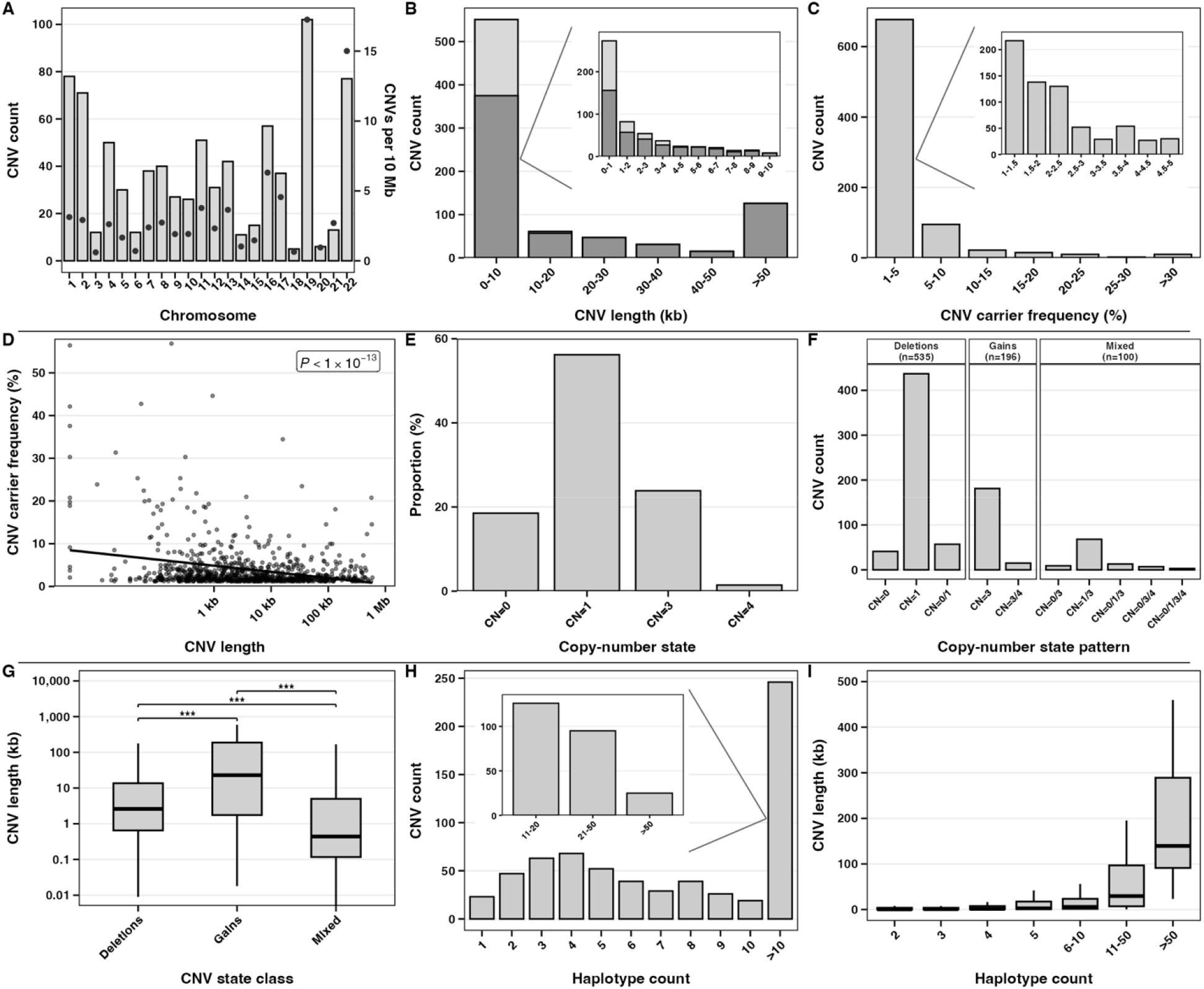
Summary of CNV landscape. (A) Bar plot showing distribution of CNVs retained for association testing across autosomes. Bars show CNV counts per chromosome, and points show CNV density normalized as CNVs per 10 Mb. (B) Bar plot showing the length distribution of retained CNVs, with haplotype-assigned CNVs shown in dark grey. (C) Bar plot showing distribution of CNV carrier frequency, defined as the percentage of individuals with an altered copy number at each CNV. (D) Scatter plot showing relationship between CNV length and CNV carrier frequency. The regression was fit using log-transformed CNV length. (E) Bar plot showing distribution of total altered copy-number states across all CNV observations. (F) Faceted bar plot showing CNV-level copy-number patterns based on whether each CNV showed deletion states, gain states, or both across individuals. (G) Boxplot showing CNV length by CNV-level state class. Mixed refers to CNVs exhibiting both gains and deletions across samples. Significance was assessed using pairwise Wilcoxon rank-sum tests with BH-correction. ***: *p* < 0.001. (H) Bar plot showing distribution of distinct haplotype counts across haplotype-resolved CNVs. (I) Boxplot showing CNV length by number of observed haplotypes.

CNV carrier frequency, defined as the proportion of individuals carrying an altered copy-number state at a given CNV, ranged from 1.25% to 56.85%, with a median of 2.08% (Fig 2C). CNV carrier frequency was inversely associated with CNV length, with shorter CNVs observed in a larger proportion of individuals across the cohort (linear model, *p* = 1.74 x 10^-15^; Fig 2D). Among all variable CNV observations, hemizygous deletions (CN=1) were most common, followed by single-copy gains (CN=3) (Fig 2E). Consistent with CN=1 being the most common altered copy-number state, the most frequent CNV-level pattern was CN=1 only, and most CNVs contained only deletion states (Fig 2F).

CNV length differed significantly across CNV dosage states (deletion, gain, or mixed; Kruskal-Wallis χ² = 109.71, df = 2, *p* < 2.2 x 10^-16^; Fig 2G). Gain-only CNVs had the greatest median length (23 kb, IQR: 1.8-188.2 kb), followed by deletion-only CNVs (2.6 kb, IQR: 0.7-13.6 kb), while mixed (deletions and gains) CNVs were shortest (0.4 kb, IQR: 0.1-5 kb). Post hoc pairwise Wilcoxon rank-sum tests with Benjamini-Hochberg (BH)-correction showed significant differences between all groups (Fig 2G).

Among CNVs with successful haplotype assignment, the number of observed haplotypes per CNV ranged from 1 to 281. Four distinct haplotypes per CNV was most common, followed by three (Fig 2H). CNVs with greater haplotypic complexity were generally larger (Fig 2I). Here, the CNV landscape was characterized by predominantly short, low-frequency events, with deletions being the most common altered copy-number state.

### Characteristics of associated CNV regions

Overall, 49 replicated associations between CNVs and bacterial species, involving 47 distinct CNV regions (5.7% of tested regions), were identified (S1 Fig). Most associations were detected through haplotype-set testing rather than CNV dosage testing (34 versus 15, respectively). Associated regions had a median length of 13.6 kb, compared with 2.8 kb for non-significant regions, and a median carrier frequency of 2.08% (range, 1.25%-9.96%). Of the associated regions, 41 (87.2%) overlapped at least one gene, and collectively, there were 66 unique genes (S3 Table). Region length was associated with significance, with each doubling in length corresponding to 20% higher odds of significance (OR = 1.20, 95% CI 1.09-1.32, *p* = 1.33 x 10^-4^), whereas carrier frequency and gene count were not significant. Post hoc analyses evaluated a median of five non-reference haplotypes per associated haplotype-set.

### Microbial associations

Associated CNVs were linked to the abundance of 25 bacterial species (25 of 68 prevalent species). Of these, 17 were associated through haplotype-set testing only, three were associated with CNV dosage only, and five were associated with both. Six CNV-associated species had not previously been linked with genetically regulated gene expression (GReX) or SNPs^23,24^. Associations involved anaerobic, facultative, and aerobic species, with 22, 25, and 2 associations, respectively. By Gram stain, 29 associations involved Gram-positive species and 20 involved Gram-negative species (S4 Table). Aerotolerance and Gram-stain classifications were not statistically overrepresented among associated species. Investigation at species-level revealed *Parvimonas micra* had the largest number of associations (6), followed by *Staphylococcus cohnii* (5), and *Fusobacterium nucleatum* (5). Associations with multiple species were observed once through CNV dosage testing, with chr19:41,385,767-41,385,869 associated with *F. nucleatum* and *Klebsiella oxytoca*, and once through haplotype-set testing, with chr8:144,990,702-144,999,224 associated with *Winkia neuii* (formerly *Actinomyces neuii*) and *Streptococcus intermedius*. Although associated species were individually detected a median of 1.66% of patients, they collectively occurred 38.6% across cohorts (186 of 482 patients). The most prevalent repeatedly associated species, *Porphyromonas bennonis*, was present in 31 patients and ranked 15th among the 68 species tested. For species without replicated associations, 1,730 associations passed hierarchical false discovery rate (FDR) in one cohort or the other (S5 Table).

### Co-occurrence of associated species

Evaluating presence of repeatedly associated species, five significant positive co-occurrences and no significant negative co-occurrences were observed, where all significant co-occurrences involved anaerobic species. Four significant positive co-occurrences involved one CNV-associated species and another species not associated with CNVs: *P. bennonis* with *Finegoldia magna*, *P. bennonis* with *Anaerococcus prevotii*, *Peptostreptococcus anaerobius* with *Prevotella disiens*, and *Peptoniphilus asaccharolyticus* with *Anaerococcus hydrogenalis* (Table 2). A notable positive co-occurrence emerged between two CNV-associated species: *Fusobacterium canifelinum* with *F. nucleatum.* Because statistically co-occurring *F. canifelinum* and *F. nucleatum* belong to the same genus, the 16S V1-V2 sequences in the clinical database representing these specific epithets were checked by sequence alignment and p-distance calculation; between 6 *F. canifelinum* and 49 *F. nucleatum* the median sequence identity was 96.44% (11 mismatches), with the most similar being 99.35 % (2 mismatches). Thus, these species assignments are plausibly correct but could also be explained by sequencing error or other technical artifacts.

**Table 2:** Significant co-occurrences involving CNV-associated wound species.

| Species 1 | Species 1 occurrence | Species 2 | Species 2 occurrence | Observed co-occurrence | Expected co-occurrence | FDR |
| --- | --- | --- | --- | --- | --- | --- |
| *Fusobacterium canifelinum* | 5 | *Fusobacterium nucleatum* | 21 | 4 | 0.2 | 0.0046 |
| *Porphyromonas bennonis* | 31 | *Finegoldia magna* | 114 | 19 | 7.3 | <0.0001 |
| *Porphyromonas bennonis* | 31 | *Anaerococcus prevotii* | 27 | 9 | 1.7 | 0.0046 |
| *Peptostreptococcus anaerobius* | 18 | *Prevotella disiens* | 13 | 5 | 0.5 | 0.0138 |
| *Peptoniphilus asaccharolyticus* | 7 | *Anaerococcus hydrogenalis* | 24 | 4 | 0.3 | 0.0413 |

### Overrepresentation analysis

To elucidate the potential functional consequences of associated CNVs, Reactome overrepresentation analysis identified 15 enriched pathways belonging to 5 parent pathways. Enriched pathways were assigned to ECM (8), sensory perception (3), developmental biology (2), metabolism (1), and signal transduction (1; Fig 3A; S6 Table). Collectively, species-specific associations were enriched for 18 Reactome pathways, belonging to 7 parent pathways. *K. oxytoca* had the greatest number of species-specific enrichments (6), followed by *Pasteurella canis* (5), *Staphylococcus haemolyticus* (3), *S. cohnii* (3), and *Serratia marcescens* (1). Sensory perception and metabolism were the parent categories enriched across multiple species (Fig 3B; S7 Table).

**Fig 3.**
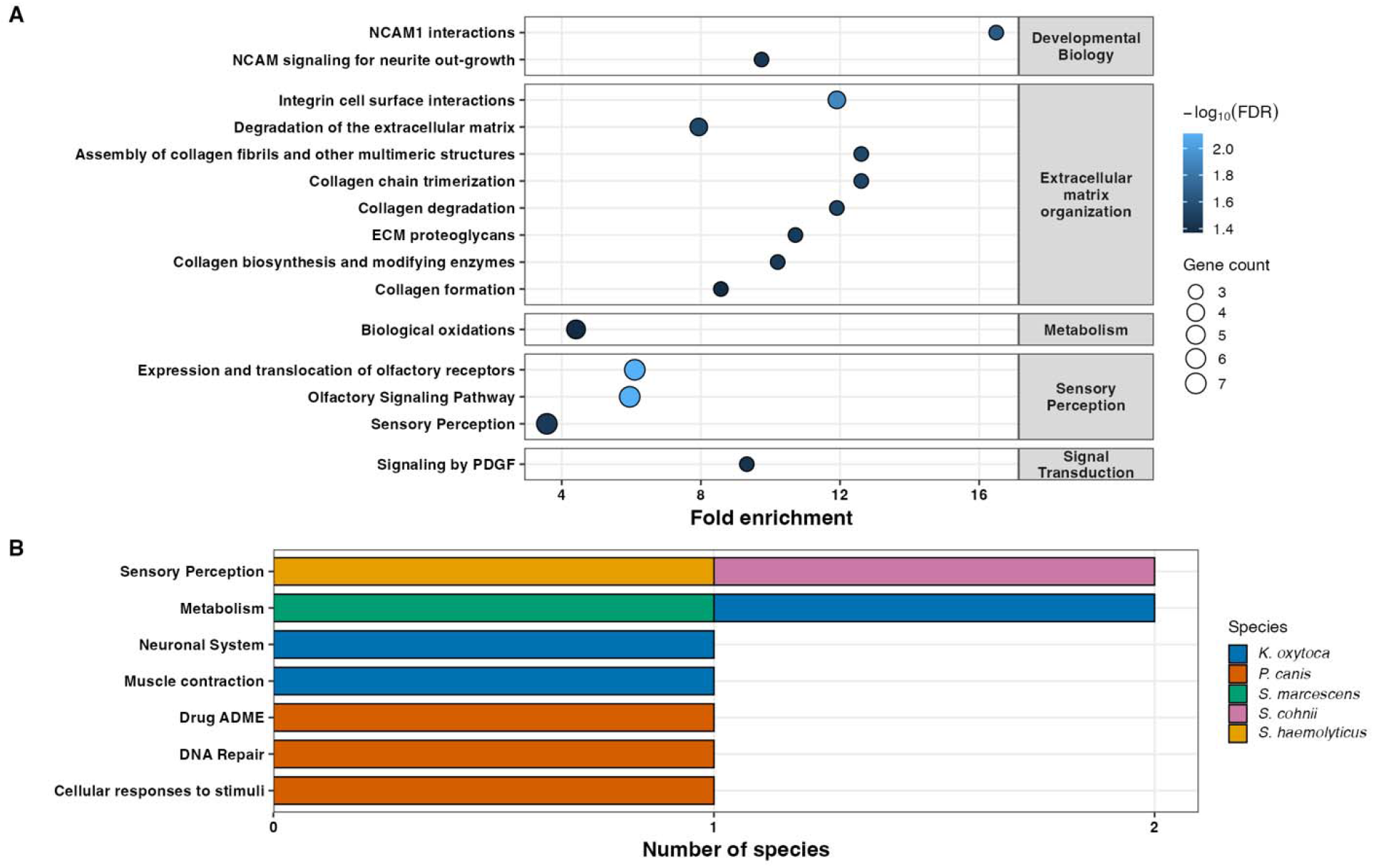
Reactome significant enrichment results for genes within associated CNVs. (A) Reactome enrichment dot plot across all CNV gene sets. Rows show enriched Reactome pathways grouped by parent Reactome pathway, shown in the grey facets on the right. Dot size represents the number of genes contributing to each pathway. (B) Number of species showing significant enrichment within each parent Reactome pathway. Bars are stacked by species, with each segment representing one contributing species.

### Additive effects models of species abundance

A total of 54 species had at least one association with a CNV, or previously with a GReX predictor^24^, or SNP^23^ and were included in additive genomic modeling. Correlated microbial phenotypes were controlled in these models with a median of four PCs (S2 Fig). The highest heritability (*h*^2^*\**) observed was 43.64% for *Anaerococcus obesiensis*. Across species, the median *h*^2^*\** was 12.84% (IQR: 6.91% - 22.58%) (Fig 4), and 30 species had 95% confidence intervals that did not span 0. The number of predictors was positively associated with *h*^2^*\**, with each additional predictor corresponding to a 0.35-percentage-point increase in *h*^2^*\** (*p* = 9.56 x 10^-6^) (S3 Fig). GReX explained the largest median *h*^2^*\** (13.15%), followed by CNVs (8.76%), and SNPs (2.78%).

**Fig 4.**
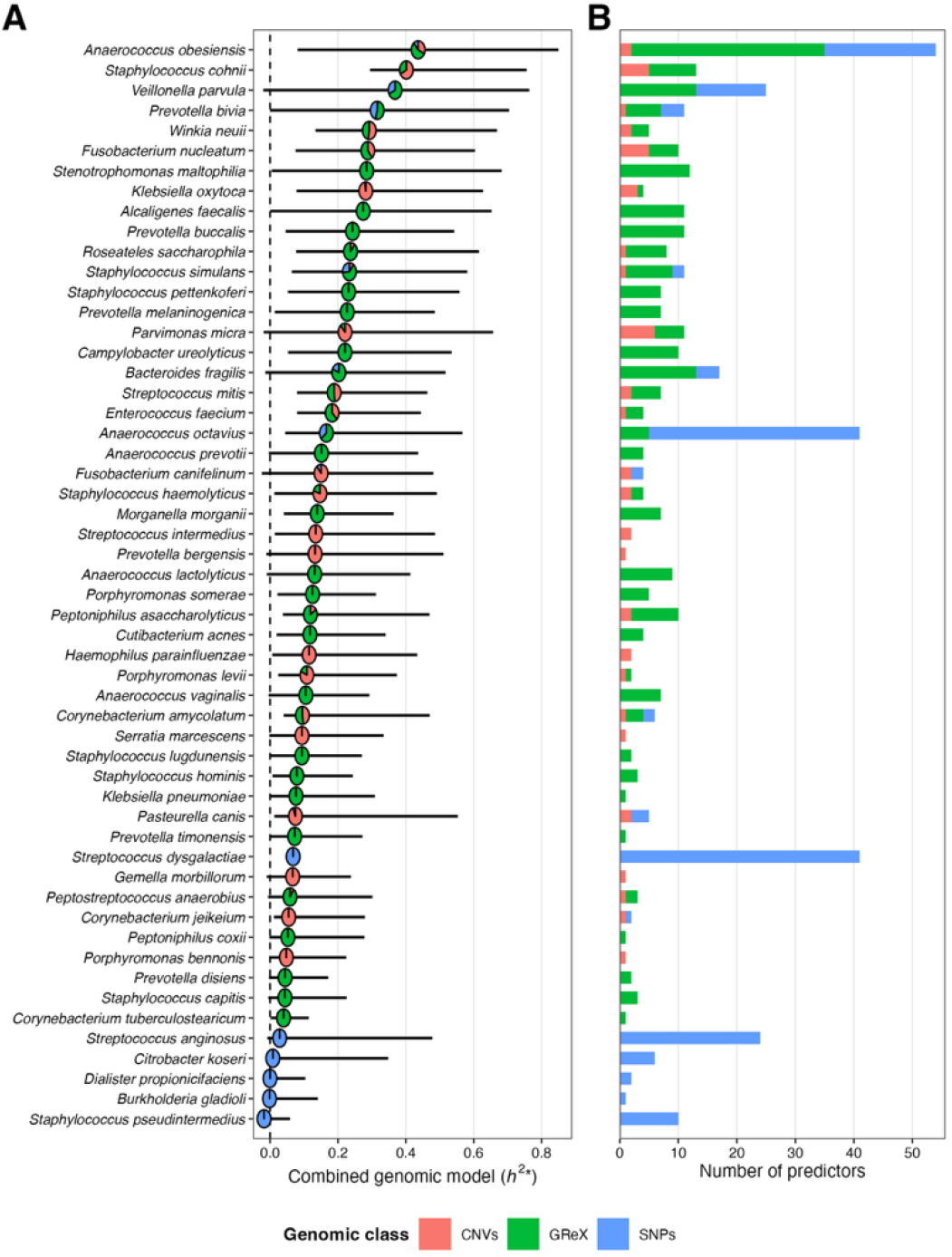
Additive genomic modeling of residual microbial species abundance. (A) Dot plot showing *h*^2^*\** (* indicates epistatic effects in CNVs and possible additive) from combined genomic models for 54 species. Horizontal lines show 95% confidence intervals. Pie slices within each point show the relative contributions of CNVs, GReX, and SNPs predictor classes, calculated as each semi-partial R^2^. Species are ordered by *h*^2^*\**. (B) Stacked bar plot showing the number of CNVs, GReX, and SNP loci included in each model.

## Discussion

The role of host factors in shaping chronic wound microbiome composition is increasingly appreciated but remains incompletely defined. Recent studies have shown that patient genetic variation, including GReX ^24^ and SNPs^23^, is associated with wound microbial diversity. Here, we extend this framework by demonstrating that CNVs represent an additional type of host genomic variation associated with the chronic wound microbiome. Using a two-cohort discovery and replication design with stringent multiple-testing control, we identified replicated associations between CNVs and bacterial species abundance. In addition to modeling CNV dosage, we inferred haplotypic variation within CNV regions and assigned haplotype dosages to evaluate whether local allelic structure provided additional resolution beyond overall copy number. Notably, when haplotype models were available, they outperformed CNV dosage models, indicating underlying allelic diversity is influential to microbial selection beyond CNV dosage alone.

The CNV landscape was dominated by short, low-frequency events and showed heterogeneity in chromosomal distribution, size, carrier frequency, copy-number state, and haplotypic complexity. The inverse relationship between CNV carrier frequency and length is consistent with population-scale studies showing that shorter CNVs are more common than larger events^37^. CNV density differed significantly across autosomes, in agreement with the established view that CNVs are unevenly distributed among chromosomes rather than occurring in direct proportion to chromosome size^38,39^. Deletions were more common than gains, consistent with previous large-scale CNV studies^31,40^, although this pattern may reflect the greater sensitivity of array-based methods for detecting deletions than duplications^41^. Gain-only CNVs were also significantly larger than deletion-only CNVs, consistent with previous work^42,43^. Here, haplotype complexity increased with CNV size, likely because of increased length and larger CNVs contained more SNPs (Spearman’s rho = 0.277, *p* = 2.44 x 10^-16^) from which distinct haplotypes could be defined. Larger CNV regions were more likely to be significant, with each doubling in length corresponding to 20% higher odds of significance. CNV size may contribute to functional impact, as pathogenic CNVs tend to be substantially longer than benign CNVs and larger CNVs overlap more functional and dosage sensitive regions^44^. Collectively, results support CNV size as an important genomic attribute influencing functional effects selecting microbial species.

Associations were detected for 25 bacterial species, and each CNV was typically associated with a single species. This pattern supports previous work^23,24^ and indicates substantial diversity in how the wound microenvironment selects species. Whereas individual species detected in the wound typically do not have large cohort-wide prevalence rates, those associated with the highest confidence were collectively detected in 38.6% of patients. Many of these species have been reported in wound infections, including the ESKAPE group member *Enterococcus faecium* ^45^ and opportunistic pathogens, *K. oxytoca*^46^, *S. haemolyticus*^47^, and *S. marcescens*^48^. Notably, *P. micra*, *S. cohnii*, and *F. nucleatum*, the species with the greatest numbers of associations, have each been documented in infected wounds^49–51^. Sequencing-based studies further demonstrate the occurrence of many of the taxa identified here in chronic wounds, including *F*. *nucleatum*, *Prevotella bivia*, and *Corynebacterium jeikeium* at the species level, with genus-level overlap including *Anaerococcus*, *Peptoniphilus*, *Peptostreptococcus*, *Porphyromonas*, and *Prevotella*^52^.

Although hallmark wound pathogens such as *Staphylococcus aureus* and *Pseudomonas aeruginosa* did not replicate across both cohorts, these and several more species were among those 1,730 additional associations passing study-wide threshold in one cohort or the other. We note that a two-cohort design, both requiring study-wide significance thresholding is quite stringent, particularly given the low carrier frequencies of CNVs and the resulting limited statistical power, as frequency is an important determinant of statistical power^53^. For example, although *S. aureus* was detected in 163 patients, the median CNV was carried by 10 patients or five carriers when split evenly across two cohorts. Therefore, associations passing both cohorts should be considered highest confidence and those passing one cohort as plausible.

Importantly, however, detection of these organisms does not necessarily indicate clinical infection. Longitudinal profiling showed that recognized pathogens, such as *S*. *aureus* and *P*. *aeruginosa* were present in clinically uninfected wounds, while infection onset was associated with decreased pathogen abundance and increased obligate anaerobes, including *Anaerococcus*, *Peptoniphilus*, *Porphyromonas*, and *Prevotella*^54^. Notably, each of these genera was represented among the CNV-associated species identified here, although obligate anaerobes were not significantly overrepresented overall. Conversely, the presence of bacteria in chronic wounds is not necessarily detrimental, as *Alcaligenes faecalis*, shown here to have significant heritability, has been experimentally shown to accelerate diabetic wound healing by promoting reepithelialization^55^. Thus, the relationship between the abundances of CNV-associated species and wound healing remains to be determined.

Because ecological interactions among wound microbial species can influence observed associations with host phenotypes, species co-occurrence patterns should be considered alongside host genetic association signals when interpreting results. Here, significant co-occurrences were always positive and involved obligate anaerobic species. Similar co-occurrences have been reported in chronic wounds, including *Porphyromonas* with *Finegoldia* and *Anaerococcus*, and *Peptoniphilus* with *Anaerococcus*^56^. Also, all but one significant co-occurrence included a CNV-associated and a non-associated species. A plausible interpretation of this pattern is host genetics promoted colonization of the selected species, which in turn modified the wound environment subsequently promoting colonization by the non-associated species. Indeed, wound-associated obligate anaerobes can be clinically relevant, producing multi-species biofilms and exhibiting antibiotic tolerance contributing to treatment-recalcitrant infections^57^.

Two CNVs were of particular interest because each was associated with multiple species. The 8,523-bp CNV at chr8:144,990,702-144,999,224 lies entirely within *PLEC* and overlaps exons in two transcripts. This region showed replicated haplotype-set associations with *W. neuii* and *S. intermedius*. PLEC is known to organize epithelial cytoskeletal networks and supports intercellular junctions, cell cohesion, and mechanical stability^58^. Its deficiency has been associated with impaired epithelial barrier function and increased bacterial penetration^59^. Reinforcing a role for *PLEC* in shaping the wound microbiome, *PLEC* overlapped two additional CNVs each independently associated with *Corynebacterium amycolatum* and *Streptococcus mitis*. Consistent with a prevalent role for cytoskeletal dynamics, previous work identified *SYNPO*, an actin-associated protein that regulates cytoskeletal organization and epithelial junctions, expression associated with *A. obesiensis* across six tissues^24^. Moreover, Tipton et al. (2020)^22^ found variation linked to the expression of *TLN2*, a mandatory mechanosensory protein linking integrins to the actin cytoskeleton, as significantly associated with microbiome variation.

The second CNV associating to multiple species was the 103-bp CNV at chr19:41,385,767-41,385,869 positioned entirely within an intron of *CYP2A7*. This region showed replicated CNV dosage associations with *F. nucleatum* and *K. oxytoca*, and increased dosage was associated with greater abundance of both species. *CYP2A7* may influence the expression of the related cytochrome P450 enzyme CYP2A6^60^, whose activity can be altered during infection and inflammation^61^. However, *CYP2A6* is predominantly expressed in the liver, and its relevance to bacterial abundance remains unclear.

Among the 49 CNV associations, collagen and ECM were repeatedly represented. Enrichment of ECM-related processes included collagen formation, biosynthesis, degradation, fibril assembly, and integrin interactions. These pathways showed 8- to 13-fold enrichment and were driven by the collagen genes *COL3A1*, *COL4A3*, and *COL6A2*, with *BSG* additionally contributing to integrin cell-surface interactions and ECM degradation pathways. Importantly, the CNVs containing collagen and *BSG* genes were not co-associated due to linkage; they were identified in four CNVs distributed across three chromosomes, and *COL3A1* and *COL4A3* are separated on chromosome 2 by more than 38 Mb. ECM components, including collagen specifically, are known substrates for bacterial adhesion and can facilitate subsequent host colonization, invasion, and dissemination^62^. *In vitro* collagen chronic wound models further demonstrate this interaction with bacteria observed to be embedded within the ECM and interacting with collagen fibers^63^.

Variation in the ECM landscape appears to select microbiomes in diverse ways. Specifically, haplotype variation at *COL3A1* and *COL4A3* was associated with *Staphylococcus simulans* and *Prevotella bergensis* abundance, respectively, while greater dosage of a specific haplotype at *COL6A2* was associated with lower *F*. *nucleatum* abundance, and greater overall *BSG* dosage was associated with lower *S. cohnii* abundance. Moreover, ECM-related functions were enriched in all previous wound microbiome association studies to date, which collectively investigated a range of types of genetic variation. For example, the afore-mentioned *TLN2* association^22^ is relevant, as *TLN2* mediates inside-out integrin activation and links activated integrins to the actin cytoskeleton, thereby facilitating cell–ECM adhesion. Omeir et al.^24^ reported expression associations with two additional collagen genes; decreased *COL9A3* expression was associated with higher *A. faecalis* abundance, and decreased *COL10A1* expression was associated with higher *Staphylococcus capitis* abundance. Gabrilska et al. (2025)^23^ additionally reported the association of *COL18A1-AS1* exonic SNPs with *Streptococcus anginosus*. Although these associations may partly reflect species-specific substrate preferences, this interpretation alone may be overly simplistic. Notably, *COL3A1, COL4A3,* and *COL6A2* also drove enrichment of NCAM1 interactions, the pathway exhibiting the greatest fold enrichment. NCAM1-mediated interactions couple the extracellular environment to processes regulating cell adhesion, migration, proliferation, differentiation, and survival, suggesting that ECM variation may influence microbial selection not only by altering available attachment substrates, but also by modifying host-cell behavior and the resulting wound microenvironment.

Host glycosylation and olfactory signaling represent other mechanisms of microbial selection. Two partially overlapping CNVs within *ABO* on chromosome 9 were associated with *F*. *nucleatum* and *S. cohnii*. *ABO* variation determines the addition of specific terminal sugars to H antigen, and consequently, ABO blood group. These antigens can influence infection by serving as receptors for bacteria or their products, facilitating colonization, invasion, or host evasion^64^. *ABO* has also been associated with both gut^65^ and oral^66^ microbiomes, supporting a broader influence of ABO on the host-microbe interface. Olfactory signaling was enriched through seven olfactory receptor family 4 genes, *OR4K3*, *OR4N2*, *OR4K2*, *OR4Q3*, *OR4M1*, *OR4K5*, and *OR4K1*, located across two distinct regions on chromosome 14. Olfactory receptors can interact with microbial metabolites and modulate inflammation, although the functions and ligands of these specific OR4 receptors remain unknown^67^. At the species level, only two pathways were enriched across more than one species: olfactory signaling across two *Staphylococcus* species (*S. haemolyticus* and *S. cohnii*) and phase I functionalization of compounds across *S. marcescens* and *K. oxytoca* through *AADAC* and *CYP2A7*, respectively. *AADAC* regulates lipid, xenobiotic, and drug metabolism and has been implicated in disorders, including obesity, diabetes, and atherosclerosis^68^. As diabetes (yes/no) was controlled as a covariate, the association of *AADAC* should be independent of any binary effects the gene may have on diabetes that in turn influences *S. marcescens* in the wound bed.

Heritability models integrating CNVs, GReX, and SNPs explained substantial microbial abundance variation across several species, with 24 species having median estimates greater than 20% and confidence intervals for 30 species did not span zero. Because models controlled for microbial correlation, in addition to patient level covariates, *h*^2^*\** should largely reflect genomic effects separated from ecological effects and the patient context more broadly. Moreover, models containing more genomic predictors typically explained greater microbial variation and given the relatively modest sample size of the current study, heritability estimates should be considered minimum. Incorporating multiple types of genetic variants showed that GReX and CNV predictors explained approximately 4.7 and 3.2 times more variation, respectively, as compared to SNP predictors, likely reflecting the broader biological information captured by these predictor classes.

This study provides robust CNV associations with the chronic wound microbiome, extending evidence for patient genetic control of wound microbial composition to structural and haplotypic variation. In addition to replicated associations, significant functional enrichment serves as an additional layer of biological validation. Moreover, consistency with, and expansion of, previous work regarding functional implications, particularly the recurrent involvement of cytoskeletal dynamics, collagen, and other ECM components, provides convergent support for a causal model in which patient genetic variation modifies the ECM and cell-ECM interactions that, in turn, shape the wound microbiome. Heritability models further showed that multiple forms of host genetic variation collectively accounted for substantial variation in microbial abundance, with GReX and CNV predictors explaining considerably more variation than SNP predictors. Regarding magnitude, significant heritabilities were calculated for 30 species, and most of these were greater than 20%, a result indicating the impact of patient genetic variation on the wound microbiome is appreciable. Larger, multicenter cohorts incorporating longitudinal microbiome sampling, clinical variables collected throughout treatment, and outcome data will be important not only for expanding our understanding of the mechanisms underlying genome-wound microbiome relationships, but also for determining how patient genetics and the microbiome interact, directly and indirectly, to influence healing and ultimately inform patient-specific treatment. For example, knowing a patient’s predisposition to specific microbes or healing trajectories could signal a need for specialized intervention.

## Methods

### Sample collection

This study made use of chronic wound patient data previously reported^22–24^. Study samples were obtained from adults who provided written informed consent under Western Institutional Review Board (IRB) protocol 56-RW-156DNA. All samples were collected in a deidentified manner. Participants were recruited at their first clinical visit to the Southwest Regional Wound Care Center (Lubbock, Texas, USA), presenting with a non-healing wound persisting for more than 3 weeks. Following clinical evaluation and treatment planning, wounds were sharply debrided to remove slough and devitalized tissue until a sanguineous wound bed was achieved. The collected debrided tissue was homogenized manually with a curette, placed into a collection tube, and transported the same day at room temperature to MicroGenDX (Lubbock, TX, USA), a CAP-accredited and CLIA-licensed high-complexity diagnostic laboratory. Buccal swabs were also collected for host DNA analysis and were immediately stored in liquid nitrogen before accessioning into the Wolcott Wound Care Research Collection at the Museum at Texas Tech University (Lubbock, TX, USA). Debrided wound tissue was used for microbiome profiling, whereas buccal swabs were used for host genomic analysis. Individuals were excluded if they had duplicate identification numbers, unavailable electronic medical records, or missing clinical reports.

### Patient DNA extraction and quantification

Genomic DNA was isolated using Gentra Puregene Buccal Kits (Qiagen, Gaithersburg, MD, USA). Swabs were lysed overnight at 55°C, and RNase A treatment was included to remove RNA contamination. Purified DNA was eluted in 50 μL of molecular biology-grade water (GE Healthcare Life Sciences, Marlborough, MA, USA). DNA concentration was measured with a Qubit 3.0 Fluorometer (Thermo Fisher Scientific, Waltham, MA, USA), and samples were adjusted by dilution or dehydration to a final concentration of 10 to 50 ng/μL. Extracted DNA was stored at -80°C until genotyping.

### Patient genotyping and quality control

Genotyping was conducted using the Illumina Infinium High-Throughput Screening Assay protocol (Illumina, San Diego, CA, USA). Briefly, genomic DNA underwent whole-genome amplification at 37°C for 20-24 hours. Amplified DNA was then fragmented and hybridized to Illumina BeadChips at 48°C for 16-24 hours. Following hybridization, unbound and nonspecifically bound DNA was removed through washing. Hybridized primers were extended using biotin- or dinitrophenyl-labeled nucleotides, after which BeadChips were coated and processed for genotype calling. The Illumina iScan System was used to generate SNP array intensity data files (IDAT). The IDAT files are publicly available through NCBI Gene Expression Omnibus repository under accession GSE276944. Analysis of deidentified data was conducted under Texas Tech University IRB, IRB2020-955. Genotype data were processed in GenomeStudio, and the PLINK Input Report Plug-in was used to export PLINK files for marker-level and individual-level quality control. Because samples were genotyped across Illumina Infinium Global Screening Array v2 and v3, datasets were merged using SNPs shared across both array versions. Markers with missingness greater than 5% were excluded. Nonautosomal variants were excluded, and only biallelic SNPs were retained. Samples were excluded for (1) high missingness (greater than 0.09), (2) outlying heterozygosity defined as more than 3 standard deviations from the mean, (3) elevated relatedness defined by identity by descent greater than 0.1875, or (4) divergent ancestry identified by principal component analysis (PCA) implemented in R (v4.5.3) using SNPRelate^69^.

### Microbiome profiling

Bacterial relative abundances were obtained from clinical reports (MicroGenDX) generated for the treating physician. Species-level microbiome profiling was performed using V1-V2 16S rRNA gene sequencing, which is commonly informative for species-level calls^70^. Detailed workflow of microbial profiling was described previously^22,24,71^. Species with study-wide prevalence rates greater than 1% were included for association testing. Relative abundances were centered log-ratio (CLR) transformed.

### CNV calling

CNVs were called from IDAT files using B allele frequency (BAF) and log R ratio (LRR) values from shared markers within a hidden Markov model framework in PennCNV^72^. Population frequency of the B allele was generated using compile_pfb.pl, and a GC content model was computed using cal_gc_snp.pl. Genome-wide genomic waves in SNP-array signal intensity correlating with GC content can distort LRR profiles and generate spurious copy-number calls, therefore GC-based wave adjustment was applied using the GC model (-gcmodel) prior to CNV filtering^73^. CNV calls were filtered at the call level to retain regions supported by at least three markers, and at the sample level to exclude noisy samples with LRR standard deviation greater than 0.35 using filter_cnv.pl. Genes overlapping CNVs were identified using the Ensembl Homo_sapiens.GRCh37.87.gtf gene annotation file^74^.

### Haplotype assignment

Haplotype-specific CNV dosage was inferred by integrating allele-specific copy number states from BAF with phased SNP haplotypes (Fig 5). Quality-controlled SNPs within each CNV were used to define phased haplotypes and assign haplotype-specific copy number. CNVs lacking informative SNP markers or lacking haplotypic variation for a given region across all samples were retained for total copy number analyses but were excluded from haplotype-set testing. For each sample and CNV region, allele-specific copy number states were assigned by binning BAF values conditional on total copy number using predefined thresholds that allow for variation around expected allele fractions. For hemizygous deletions (CN=1), BAF values over 0.5 were classified as B and values below 0.5 as A. For single-copy gains (CN=3), A/A/A was assigned for BAF < 1/6, A/A/B for 1/6 to 1/2, A/B/B for 1/2 to 5/6, and B/B/B for BAF > 5/6. For double-copy gains (CN=4), A/A/A/A was assigned for BAF < 1/8, A/A/A/B for 1/8 to 3/8, A/A/B/B for 3/8 to 5/8, A/B/B/B for 5/8 to 7/8, and B/B/B/B for BAF > 7/8. Allelic states were converted to nucleotide-resolved compositions using Illumina manifest annotation for A/B alleles. Homozygous deletions (CN=0) were treated as having no alleles present and therefore no haplotypes assigned, whereas hemizygous deletions (CN=1) were represented by a single haplotype consistent with the observed allelic state. For diploid regions (CN=2), phased genotypes defined the two haplotypes directly. SNP genotypes were restricted to markers within CNV regions and phased using Eagle (v2.4.1) ^75^with an hg19 genetic map, producing two homologous haplotypes. For gain states (CN=3 and CN=4), BAF-inferred allelic composition at informative heterozygous SNPs was compared with phased haplotype alleles to identify the haplotype carrying the overrepresented allele. This haplotype was represented with one additional copy for CN=3 or two additional copies for CN=4 states, producing haplotype-specific copy number dosages. When all SNPs within a region were homozygous in the phased genotype data, the two haplotypes were therefore not allelically distinguishable. Such individuals were represented as carrying three or four identical haplotype copies according to their total copy number state. For each CNV region, haplotype dosage was defined as the number of copies of each inferred haplotype carried by each patient. Haplotypes present in less than 1% of patients were collapsed into a single rare-haplotype category to evaluate their collective contribution while limiting sparse predictors.

**Fig 5.**
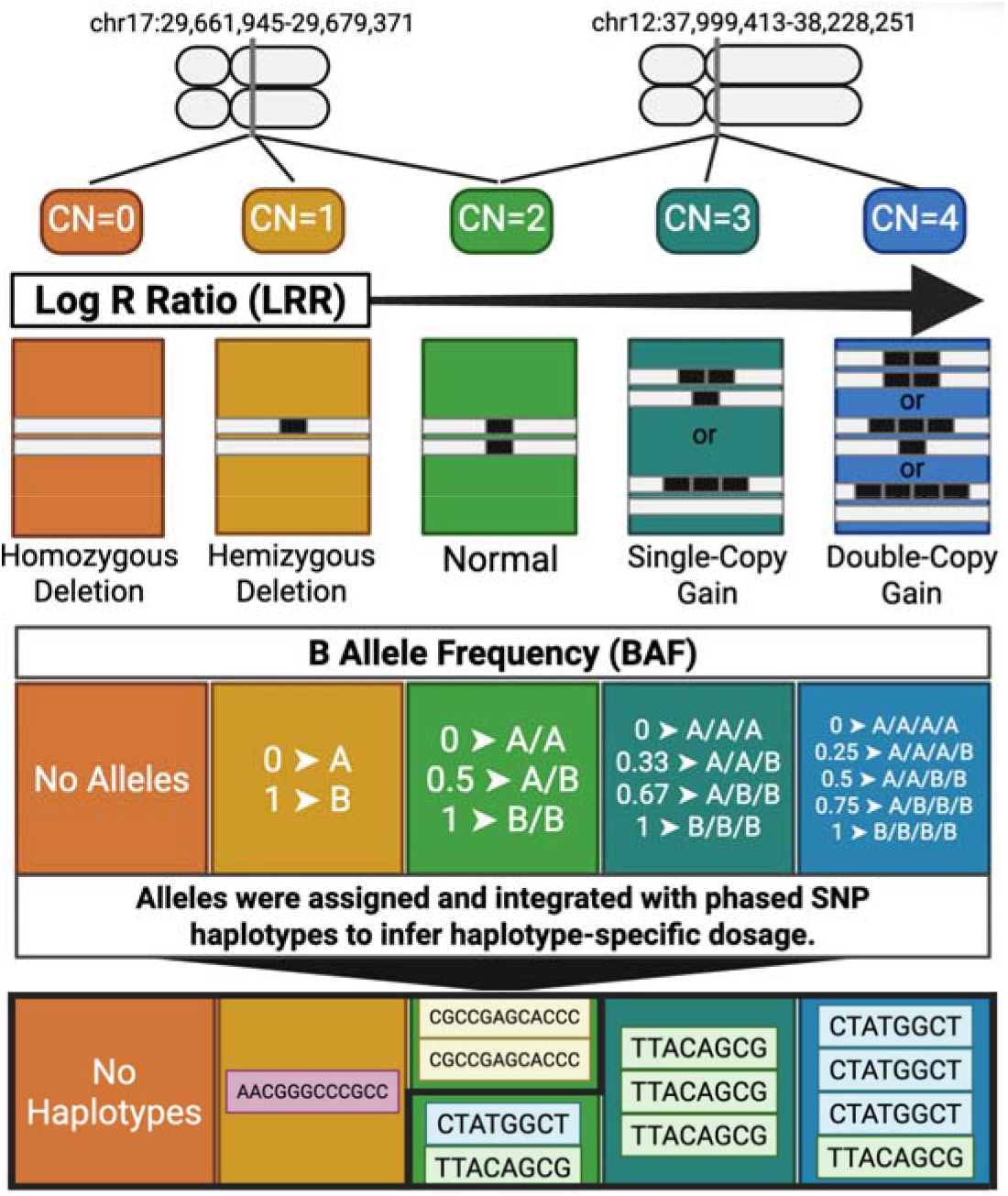
Schematic of haplotype dosage assignment. Two representative CNV regions illustrate how total CNV calls were converted into haplotype-specific dosage. On the left, the deletion-only example region, chr17:29,661,945-29,679,371, shows how CN=0 was represented by no assigned haplotypes, CN=1 by one retained haplotype copy, such as H1, and CN=2 by two haplotype copies, such as H2/H2. On the right, the gain-only example region, chr12:37,999,413-38,228,251, shows how CN=2 was represented by two haplotypes, such as H1/H2, CN=3 by three haplotype copies, such as H2/H2/H2, and CN=4 by four haplotype copies, such as H1/H1/H1/H2. Created with BioRender.com

### Cohort construction and ancestry adjustment

Samples were divided into two balanced cohorts for each tested species, stratified by self-reported race/ethnicity, species presence/absence, and normal versus non-normal copy-number status. Genetic ancestry covariates were selected by first conducting variance-standardized PCA in PLINK using the --pca option. The explanatory power of the first ten principal components (PCs) for self-reported race/ethnicity was then assessed by PERMANOVA using Euclidean distances on a binary-coded self-reported race/ethnicity matrix. Prior work has shown that, for K distinct populations, population-informative variation is captured primarily by the largest K − 1 PCs^76,77^. Because patients in this study belonged to three self-reported race/ethnicity groups, White, Hispanic, and Black, consistent with the demographics of Lubbock, TX, the main between-group ancestry structure was expected to be concentrated in the first two PCs. PC1 and PC2 were significant and explained approximately 86% of race/ethnicity-associated variance, whereas PC3-10 were not significant and cumulatively explained only 0.2% of additional race/ethnicity-associated variation. Additional PCs were not included because they did not explain meaningful race/ethnicity-associated structure in this cohort, and including PCs that do not account for population structure can imprecisely add noise and reduce power^78^.

### CNV dosage and haplotype-set association testing

For each microbial species, CNV region, and cohort, two linear models were fitted using CLR-transformed species abundance while adjusting for diabetic diagnosis, age, sex, and PC1 and PC2. The first model included total copy number only, and, for regions with resolved haplotype dosage, the second included total copy number and haplotype dosages jointly. To avoid multicollinearity, the most prevalent haplotype across both cohorts was omitted as the reference for each CNV. Haplotype contribution beyond total copy number was evaluated by comparing the two models with an omnibus F-test. For each cohort, study-wide significance was assessed using a hierarchical FDR framework^79^. Per-species association p-values were first combined separately for CNV dosage and haplotype-set associations using Simes’ method ^80^ to generate one p-value per tested CNV region for each association class. The resulting CNV dosage and haplotype-set p-values were then adjusted across all tested regions using the BH procedure^81^. For regions passing FDR < 0.05, per-species p-values were BH-adjusted within each selected region and evaluated against a threshold of 0.05 x number of selected regions / total number of tested regions. A CNV dosage association was considered replicated when the association passed this procedure in both cohorts and had concordant beta coefficient signs. A haplotype-set association was considered replicated when the association passed the hierarchical FDR procedure in both cohorts based on the omnibus p-value. To evaluate whether CNV characteristics influenced significance, logistic regression modeled significance using log2-transformed length, gene count, and carrier frequency as predictors.

### Co-occurrence analysis

Given species are treated as phenotypes but species interactions can alter their abundances, pairwise species co-occurrences were evaluated using the cooccur package^36^. Co-occurrence was assessed using species presence/absence data, after which pairs involving at least one CNV-associated species were retained. Positive and negative co-occurrence probabilities were adjusted for multiple testing using the BH-procedure across all retained species pairs, and significance was defined by FDR < 0.05.

### Phenotypic enrichment of associated species

To assess if broad phenotypic classifications were overrepresented among significant associations, species were grouped by Gram stain and aerotolerance. Binomial tests were implemented using the total number of significant associations as the number of trials, the number of significant associations involving each group as the number of successes, and the proportion of tested species assigned to each group as the expected probability of success. Multiple-testing correction was applied using the BH-procedure separately for Gram stain and aerotolerance tests.

### Overrepresentation analysis

Reactome pathway overrepresentation analysis was performed for genes within significant CNVs in R (v4.5.3) using ReactomePA^82^. Genes within all called CNVs were used as the background gene set. Enrichment analyses were performed overall and after grouping associations by species. Only species with at least three associated genes were considered. Significant enrichment was defined as BH-adjusted FDR < 0.05. To meet ReactomePA input requirements, gene identifiers were converted to ENTREZ IDs using the bitr() function from clusterProfiler^83^. Enriched Reactome pathways were further assigned to parent pathway categories using ReactomeContentService4R^84^.

### Heritability

To evaluate the contribution of host genomic variation to chronic wound microbial species abundance, additive genomic effects on species abundance were assessed using covariate- and microbiome-adjusted residual regressions. Species with at least one identified host genomic association were considered. To account for clinical covariates, ancestry, and correlation among microbial phenotypes, each species was adjusted for diabetic diagnosis, age, sex, genomic PC1 and PC2, as well as microbial PCs. Microbial PCs were used to account for correlation among microbial phenotypes and were calculated from all other CLR-transformed species abundances after those species were individually adjusted for diabetic diagnosis, age, sex, PC1, and PC2. The number of microbial PCs included was selected separately for each species using a scree plot elbow rule. Residuals from the species model represented the portion of species abundance variation not explained by clinical covariates, ancestry, or microbial correlation structure. These residuals were subsequently modeled as the dependent variable in additive genomic regressions containing all available species-associated predictors. Predictor classes included CNV dosages and individual haplotype dosages identified in the present study, and previously reported GReX predictors ^24^ and SNP dosages^23^. Regressions were fitted using lm() in R (v4.5.3). *h*^2^* was defined as the adjusted R^2^ of each genomic model, representing the proportion of residual species abundance variation explained solely by genomic predictors. Note, the asterisk *h*^2^*\** denotes that because CNVs may include epistatic effects in addition to additive effects, reported heritability may not be strictly narrow sense as denoted by *h*^2^. Uncertainty in *h*^2^* was evaluated using 10,000 bootstrap iterations. In each iteration, resampling with replacement was performed, the genomic model was refitted, and *h*^2^* was estimated. 95% confidence intervals were calculated from the resulting *h*^2^* distributions.

## Data availability

The genomic data used for this study is available in the Gene Expression Omnibus repository under accession number GSE276944 and can be accessed using the following URL: https://www.ncbi.nlm.nih.gov/geo/query/acc.cgi?acc=GSE276944.

## Acknowledgements

We thank MicroGenDX and its CEO, Rick Martin, for their support of this work. We acknowledge the staff at the Southwest Regional Wound Care Center for their commitment to excellence in wound care, scientific collaboration, and logistical support. We are grateful to the Natural Science Research Laboratory at Texas Tech University for facilitating sample storage and long-term preservation.

## Supporting information

**S1 Table. Characteristics of 68 tested species.**

**S2 Table. Characteristics of 831 tested CNVs.**

**S3 Table. Genes associated with 47 significant CNVs.**

**S4 Table. Details of 49 replicated associations across 25 species.**

**S5 Table. 1,730 one cohort associations with species lacking replicated associations.**

**S6 Table. 15 significant Reactome pathways across all species collectively.**

**S7 Table. 18 significant Reactome pathways for species-specific associations.**

**S1 Fig: Ideogram of non-significant and significant CNVs across autosomes.** Grey circles indicate non-significant CNVs, and purple squares indicate significant CNVs identified as significant in two-cohort testing. Chromosome shading represents gene density.

**S2 Fig. Species-specific microbial principal component scree plots.** Scree plots show the proportion of variance explained by the first 15 microbial principal components for each species. The enlarged point and dashed vertical line indicate the number of microbial principal components selected for adjustment in the residual abundance model.

**S3 Fig: Regression plot illustrating the relationship between *h2\** (* indicates epistatic effects in CNVs in addition to additive effects are possible) and the number of predictors from significant associations with each species.** Points represent individual species, and the line shows the fitted linear regression. The shaded band represents the 95% CI. Some points are not labeled to reduce clutter.

## References

1 Kyaw, B. M. et al. Need for Improved Definition of “Chronic Wounds” in Clinical Studies. Acta Derm Venereol 98, 157–158 (2018). 10.2340/00015555-2786

2 Falanga, V. et al. Chronic wounds. Nat Rev Dis Primers 8, 50 (2022). 10.1038/s41572-022-00377-3

3 Wolcott, R. D. et al. Analysis of the chronic wound microbiota of 2,963 patients by 16S rDNA pyrosequencing. Wound Repair Regen 24, 163–174 (2016). 10.1111/wrr.12370

4 Fife, C. E., Eckert, K. A. & Carter, M. J. Publicly Reported Wound Healing Rates: The Fantasy and the Reality. Adv Wound Care (New Rochelle) 7, 77–94 (2018). 10.1089/wound.2017.0743

5 Lavery, L. A. et al. WHS (Wound Healing Society) guidelines update: Diabetic foot ulcer treatment guidelines. Wound Repair Regen 32, 34–46 (2024). 10.1111/wrr.13133

6 Abdelhakim, M. & Ogawa, R. Emerging Therapies in Chronic Wound Healing: Advances in Stem Cell Therapy, Growth Factor Modulation, Mechanical Strategies and Adjuvant Interventions. Dermatol Ther (Heidelb) 15, 3533–3545 (2025). 10.1007/s13555-025-01564-2

7 Carter, M. J. et al. Chronic wound prevalence and the associated cost of treatment in Medicare beneficiaries: changes between 2014 and 2019. J Med Econ 26, 894–901 (2023). 10.1080/13696998.2023.2232256

8 Queen, D. & Harding, K. in Int Wound J Vol. 21 e14709 (2024).

9 Mahmoudi, M. & Gould, L. J. Opportunities and Challenges of the Management of Chronic Wounds: A Multidisciplinary Viewpoint. Chronic Wound Care Management and Research 7, 27–36 (2020). 10.2147/CWCMR.S260136

10 Beyene, R. T., Derryberry, S. L., Jr. & Barbul, A. The Effect of Comorbidities on Wound Healing. Surg Clin North Am 100, 695–705 (2020). 10.1016/j.suc.2020.05.002

11 Xu, S. et al. The Risk of and Associated Demographic and Laboratory Variables for Amputations for Inpatients with Diabetic Foot Ulcers. Endocr Pract 30, 758–764 (2024). 10.1016/j.eprac.2024.04.019

12 Cascini, S. et al. Survival and factors predicting mortality after major and minor lower-extremity amputations among patients with diabetes: a population-based study using health information systems. BMJ Open Diabetes Res Care 8 (2020). 10.1136/bmjdrc-2020-001355

13 Uberoi, A., McCready-Vangi, A. & Grice, E. A. The wound microbiota: microbial mechanisms of impaired wound healing and infection. Nat Rev Microbiol 22, 507–521 (2024). 10.1038/s41579-024-01035-z

14 Gould, L. J. et al. WHS guidelines for the treatment of pressure ulcers—2023 update. Wound Repair and Regeneration 32, 6–33 (2024). 10.1111/wrr.13130

15 Idensohn, P. et al. International guideline on antimicrobial stewardship and the role of microbial-binding dressings in wound care 2026: infection prevention, control, early intervention and treatment. J Wound Care 35, S1–s40 (2026). 10.12968/jowc.2026.0302

16 Clemett, V. J. Bioburden and wound infections: reduction, identification and management. Br J Nurs 35, S16–s23 (2026). 10.12968/bjon.2024.0439

17 Rhoads, D. D., Cox, S. B., Rees, E. J., Sun, Y. & Wolcott, R. D. Clinical identification of bacteria in human chronic wound infections: culturing vs. 16S ribosomal DNA sequencing. BMC Infect Dis 12, 321 (2012). 10.1186/1471-2334-12-321

18 Mahnic, A., Breznik, V., Bombek Ihan, M. & Rupnik, M. Comparison Between Cultivation and Sequencing Based Approaches for Microbiota Analysis in Swabs and Biopsies of Chronic Wounds. Front Med (Lausanne) 8, 607255 (2021). 10.3389/fmed.2021.607255

19 Dowd, S. E., Wolcott, R. D., Kennedy, J., Jones, C. & Cox, S. B. Molecular diagnostics and personalised medicine in wound care: assessment of outcomes. J Wound Care 20, 232, 234–239 (2011). 10.12968/jowc.2011.20.5.232

20 Schmidt, B. M., Kaye, K. S., Armstrong, D. G. & Pop-Busui, R. Empirical Antibiotic Therapy in Diabetic Foot Ulcer Infection Increases Hospitalization. Open Forum Infect Dis 10, ofad495 (2023). 10.1093/ofid/ofad495

21 Campbell, A. et al. The heterogeneous wound microbiome varies with wound care pain, dressing type, and inflammatory gene expression. Wound Repair Regen 32, 811–825 (2024). 10.1111/wrr.13184

22 Tipton, C. D. et al. Patient genetics is linked to chronic wound microbiome composition and healing. PLoS Pathog 16, e1008511 (2020). 10.1371/journal.ppat.1008511

23 Gabrilska, R. A. et al. Functionally enriched human polymorphisms associate to species in the chronic wound microbiome. npj Biofilms and Microbiomes 11, 194 (2025). 10.1038/s41522-025-00831-6

24 Omeir, K. et al. Heritable Tissue-Specific Gene Expression Associates With Chronic Wound Microbial Species. Wound Repair Regen 33, e70055 (2025). 10.1111/wrr.70055

25 Ancira, J. et al. A structural equation model predicts chronic wound healing time using patient characteristics and wound microbiome composition. Wound Repair Regen 33, e70004 (2025). 10.1111/wrr.70004

26 Pös, O. et al. DNA copy number variation: Main characteristics, evolutionary significance, and pathological aspects. Biomed J 44, 548–559 (2021). 10.1016/j.bj.2021.02.003

27 Pande, S., Dawood, M. & Grochowski, C. M. Structural Variants: Mechanisms, Mapping, and Interpretation in Human Genetics. Genes (Basel) 16 (2025). 10.3390/genes16080905

28 Hujoel, M. L. A. et al. Protein-altering variants at copy number-variable regions influence diverse human phenotypes. Nat Genet 56, 569–578 (2024). 10.1038/s41588-024-01684-z

29 Hastings, P. J., Lupski, J. R., Rosenberg, S. M. & Ira, G. Mechanisms of change in gene copy number. Nat Rev Genet 10, 551–564 (2009). 10.1038/nrg2593

30 Shaikh, T. H. Copy Number Variation Disorders. Curr Genet Med Rep 5, 183–190 (2017). 10.1007/s40142-017-0129-2

31 Zou, X. Z. et al. Phenome-wide analysis of copy number variants in 470,727 UK Biobank genomes. Nature 652, 675–685 (2026). 10.1038/s41586-025-10087-x

32 Poole, A. C. et al. Human Salivary Amylase Gene Copy Number Impacts Oral and Gut Microbiomes. Cell Host Microbe 25, 553–564.e557 (2019). 10.1016/j.chom.2019.03.001

33 Wang, J., Guo, H., Qi, W. & Qiao, Z. Association between Gut Microbiota Composition and Copy Number Variations in Human Genes FAM66D and TAS2R43. J Microbiol Biotechnol 35, e2504011 (2025). 10.4014/jmb.2504.04011

34 Jones, E. A., Kananurak, A., Bevins, C. L., Hollox, E. J. & Bakaletz, L. O. Copy number variation of the beta defensin gene cluster on chromosome 8p influences the bacterial microbiota within the nasopharynx of otitis-prone children. PLoS One 9, e98269 (2014). 10.1371/journal.pone.0098269

35 Fan, W. et al. Association between Human Genetic Variants and the Vaginal Bacteriome of Pregnant Women. mSystems 6, e0015821 (2021). 10.1128/mSystems.00158-21

36 Griffith, D. M., Veech, J. A. & Marsh, C. J. cooccur: Probabilistic Species Co-Occurrence Analysis in R. *Journal of Statistical Software*, Code Snippets 69, 1–17 (2016). 10.18637/jss.v069.c02

37 Durbin, R. M. et al. A map of human genome variation from population-scale sequencing. Nature 467, 1061–1073 (2010). 10.1038/nature09534

38 Veerappa, A. M. et al. Global patterns of large copy number variations in the human genome reveal complexity in chromosome organization. Genet Res (Camb) 97, e18 (2015). 10.1017/s0016672315000191

39 Zarrei, M., MacDonald, J. R., Merico, D. & Scherer, S. W. A copy number variation map of the human genome. Nature Reviews Genetics 16, 172–183 (2015). 10.1038/nrg3871

40 Kołomański, M., Szyda, J., Frąszczak, M. & Mielczarek, M. DNA sequence features underlying large-scale duplications and deletions in human. Journal of Applied Genetics 63, 527–533 (2022). 10.1007/s13353-022-00704-0

41 Balagué-Dobón, L., Cáceres, A. & González, J. R. Fully exploiting SNP arrays: a systematic review on the tools to extract underlying genomic structure. Brief Bioinform 23 (2022). 10.1093/bib/bbac043

42 Auwerx, C. et al. The individual and global impact of copy-number variants on complex human traits. Am J Hum Genet 109, 647–668 (2022). 10.1016/j.ajhg.2022.02.010

43 Stylianou, C. E. et al. Germline copy number variants and endometrial cancer risk. Hum Genet 143, 1481–1498 (2024). 10.1007/s00439-024-02707-9

44 Schuetz, R. J., Ceyhan, D., Antoniou, A. A., Chaudhari, B. P. & White, P. CNVoyant a machine learning framework for accurate and explainable copy number variant classification. Scientific Reports 14, 22411 (2024). 10.1038/s41598-024-72470-4

45 Jeyabalan, N. et al. Enterococcus faecium colonization and persistence in a model of diabetic wound infection. Infect Immun 94, e0065225 (2026). 10.1128/iai.00652-25

46 Yang, J. et al. Klebsiella oxytoca Complex: Update on Taxonomy, Antimicrobial Resistance, and Virulence. Clin Microbiol Rev 35, e0000621 (2022). 10.1128/cmr.00006-21

47 Eltwisy, H. O. et al. Pathogenesis of Staphylococcus haemolyticus on primary human skin fibroblast cells. Virulence 11, 1142–1157 (2020). 10.1080/21505594.2020.1809962

48 Boldeanu, L., Boldeanu, M. V., Novac, M. B., Assani, M. Z. & Radu, L. Serratia marcescens: A Versatile Opportunistic Pathogen with Emerging Clinical and Biotechnological Significance. Int J Mol Sci 26 (2025). 10.3390/ijms262311479

49 Heravi, F. S., Zakrzewski, M., Vickery, K., Malone, M. & Hu, H. Metatranscriptomic Analysis Reveals Active Bacterial Communities in Diabetic Foot Infections. Front Microbiol 11, 1688 (2020). 10.3389/fmicb.2020.01688

50 Kim, J. J., Lydecker, A., Davé, R., Bork, J. T. & Roghmann, M. C. Diabetic Foot Infections: Local Prevalence of and Case-Control Study of Risk Factors for Methicillin-Resistant Staphylococcus aureus and Pseudomonas aeruginosa. Open Forum Infect Dis 7, ofaa412 (2020). 10.1093/ofid/ofaa412

51 Villa, F. et al. Anaerobes in diabetic foot infections: pathophysiology, epidemiology, virulence, and management. Clin Microbiol Rev 37, e0014323 (2024). 10.1128/cmr.00143-23

52 Morsli, M. et al. Direct metagenomics investigation of non-surgical hard-to-heal wounds: a review. Ann Clin Microbiol Antimicrob 23, 39 (2024). 10.1186/s12941-024-00698-z

53 Sham, P. C. & Purcell, S. M. Statistical power and significance testing in large-scale genetic studies. Nat Rev Genet 15, 335–346 (2014). 10.1038/nrg3706

54 Radzieta, M. et al. Anaerobe-associated microbial shifts at infection onset in diabetes-related foot ulcers revealed by longitudinal metagenomics. Front Cell Infect Microbiol 16, 1812721 (2026). 10.3389/fcimb.2026.1812721

55 White, E. K. et al. Alcaligenes faecalis corrects aberrant matrix metalloproteinase expression to promote reepithelialization of diabetic wounds. Sci Adv 10, eadj2020 (2024). 10.1126/sciadv.adj2020

56 Choi, Y. et al. Co-occurrence of Anaerobes in Human Chronic Wounds. Microb Ecol 77, 808–820 (2019). 10.1007/s00248-018-1231-z

57 Coluccio, A., Lopez Palomera, F. & Spero, M. A. Anaerobic bacteria in chronic wounds: Roles in disease, infection and treatment failure. Wound Repair Regen 32, 840–857 (2024). 10.1111/wrr.13208

58 Prechova, M. et al. Plectin-mediated cytoskeletal crosstalk controls cell tension and cohesion in epithelial sheets. J Cell Biol 221 (2022). 10.1083/jcb.202105146

59 Krausova, A. et al. Plectin ensures intestinal epithelial integrity and protects colon against colitis. Mucosal Immunol 14, 691–702 (2021). 10.1038/s41385-021-00380-z

60 Nakano, M. et al. CYP2A7 Pseudogene Transcript Affects CYP2A6 Expression in Human Liver by Acting as a Decoy for miR-126*. Drug Metabolism and Disposition 43, 703–712 (2015). 10.1124/dmd.115.063255

61 Lenoir, C., Rollason, V., Desmeules, J. A. & Samer, C. F. Influence of Inflammation on Cytochromes P450 Activity in Adults: A Systematic Review of the Literature. Frontiers in Pharmacology Volume 12 - 2021 (2021). 10.3389/fphar.2021.733935

62 Wang, Y. et al. Mechanobiological mechanisms of bacterial infection-host defense interactions. Cell Reports Physical Science 7, 103347 (2026). 10.1016/j.xcrp.2026.103347

63 Price, B. L., Lovering, A. M., Bowling, F. L. & Dobson, C. B. Development of a Novel Collagen Wound Model To Simulate the Activity and Distribution of Antimicrobials in Soft Tissue during Diabetic Foot Infection. Antimicrob Agents Chemother 60, 6880–6889 (2016). 10.1128/aac.01064-16

64 Cooling, L. Blood Groups in Infection and Host Susceptibility. Clin Microbiol Rev 28, 801–870 (2015). 10.1128/cmr.00109-14

65 Rühlemann, M. C. et al. Genome-wide association study in 8,956 German individuals identifies influence of ABO histo-blood groups on gut microbiome. Nature Genetics 53, 147–155 (2021). 10.1038/s41588-020-00747-1

66 Kamitaki, N. et al. Human and bacterial genetic variation shape oral microbiomes and health. Nature 651, 429–439 (2026). 10.1038/s41586-025-10037-7

67 Orecchioni, M., Matsunami, H. & Ley, K. Olfactory receptors in macrophages and inflammation. Front Immunol 13, 1029244 (2022). 10.3389/fimmu.2022.1029244

68 Yang, L., Liao, Z. Z., Ran, L. & Xiao, X. H. Progress of arylacetamide deacetylase research in metabolic diseases. Front Oncol 15, 1564419 (2025). 10.3389/fonc.2025.1564419

69 Zheng, X. et al. A high-performance computing toolset for relatedness and principal component analysis of SNP data. Bioinformatics 28, 3326–3328 (2012). 10.1093/bioinformatics/bts606

70 Kullar, R. et al. Next-Generation Sequencing in Periprosthetic Joint Infections: Clinicians’ Guide to Its Diagnostic Role. Infectious Diseases in Clinical Practice 33, e1448 (2025). 10.1097/ipc.0000000000001448

71 Crawford, E. et al. Microbiomes in Post–Digital Rectal Exam Urine Samples are Linked to Prostate Cancer Risk. JU Open Plus 1 (2023). 10.1097/JU9.0000000000000082

72 Wang, K. et al. PennCNV: an integrated hidden Markov model designed for high-resolution copy number variation detection in whole-genome SNP genotyping data. Genome Res 17, 1665–1674 (2007). 10.1101/gr.6861907

73 Diskin, S. J. et al. Adjustment of genomic waves in signal intensities from whole-genome SNP genotyping platforms. Nucleic Acids Res 36, e126 (2008). 10.1093/nar/gkn556

74 Dyer, S. C. et al. Ensembl 2025. Nucleic Acids Res 53, D948–d957 (2025). 10.1093/nar/gkae1071

75 Loh, P. R. et al. Reference-based phasing using the Haplotype Reference Consortium panel. Nat Genet 48, 1443–1448 (2016). 10.1038/ng.3679

76 Patterson, N., Price, A. L. & Reich, D. Population Structure and Eigenanalysis. PLOS Genetics 2, e190 (2006). 10.1371/journal.pgen.0020190

77 Ma, S. & Shi, G. On rare variants in principal component analysis of population stratification. BMC Genet 21, 34 (2020). 10.1186/s12863-020-0833-x

78 Peloso, G. M. & Lunetta, K. L. Choice of population structure informative principal components for adjustment in a case-control study. BMC Genet 12, 64 (2011). 10.1186/1471-2156-12-64

79 Peterson, C. B., Bogomolov, M., Benjamini, Y. & Sabatti, C. Many Phenotypes Without Many False Discoveries: Error Controlling Strategies for Multitrait Association Studies. Genet Epidemiol 40, 45–56 (2016). 10.1002/gepi.21942

80 Simes, R. J. An improved Bonferroni procedure for multiple tests of significance. Biometrika 73, 751–754 (1986). 10.1093/biomet/73.3.751

81 Benjamini, Y. & Hochberg, Y. Controlling the False Discovery Rate: A Practical and Powerful Approach to Multiple Testing. Journal of the Royal Statistical Society. Series B (Methodological) 57, 289–300 (1995).

82 Yu, G. & He, Q. Y. ReactomePA: an R/Bioconductor package for reactome pathway analysis and visualization. Mol Biosyst 12, 477–479 (2016). 10.1039/c5mb00663e

83 Wu, T. et al. clusterProfiler 4.0: A universal enrichment tool for interpreting omics data. Innovation (Camb) 2, 100141 (2021). 10.1016/j.xinn.2021.100141

84 Poon, C.-L. et al. R interface to the reactome graph database. F1000Research 10, 721 (2021). 10.7490/f1000research.1118690.1

