## Supplementary figures and images for "The Structural Genome Shapes the Chronic Wound Microbiome"

### Supplemental Figure 1

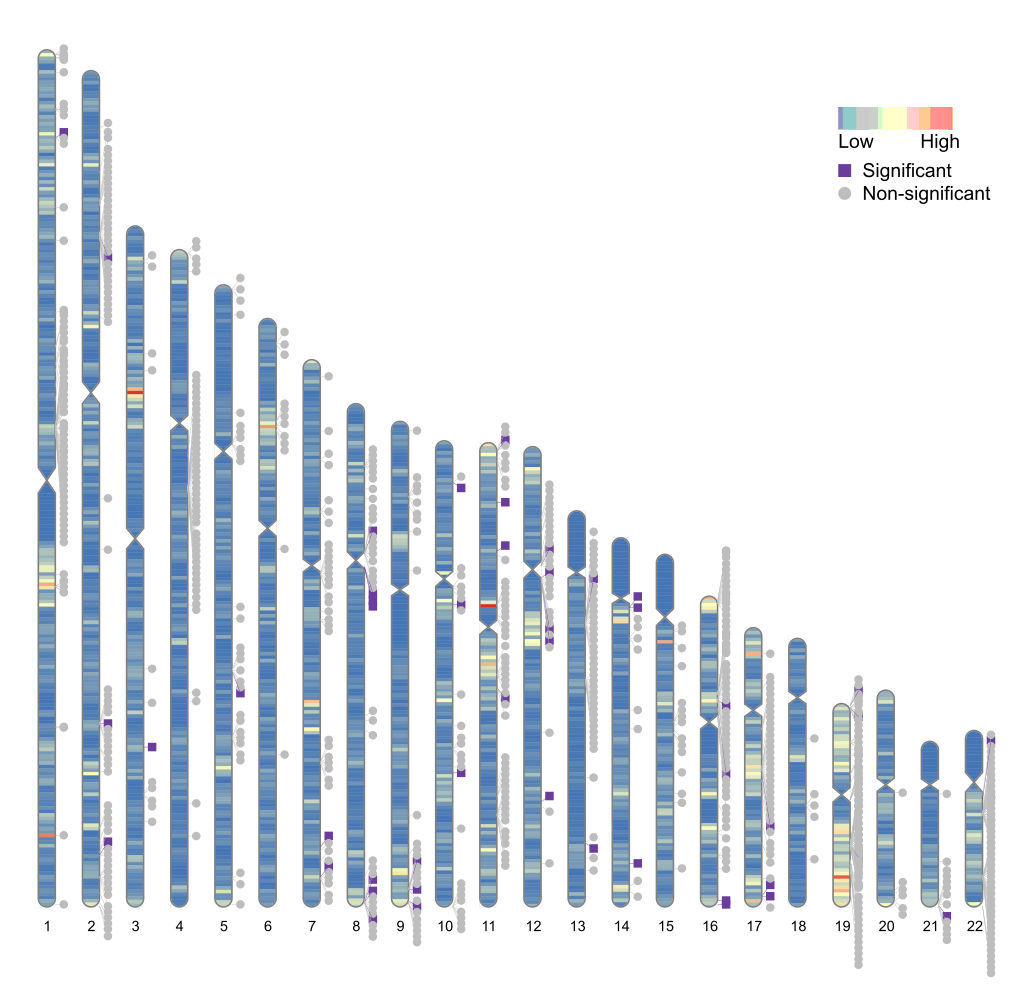

### Supplemental Figure 2

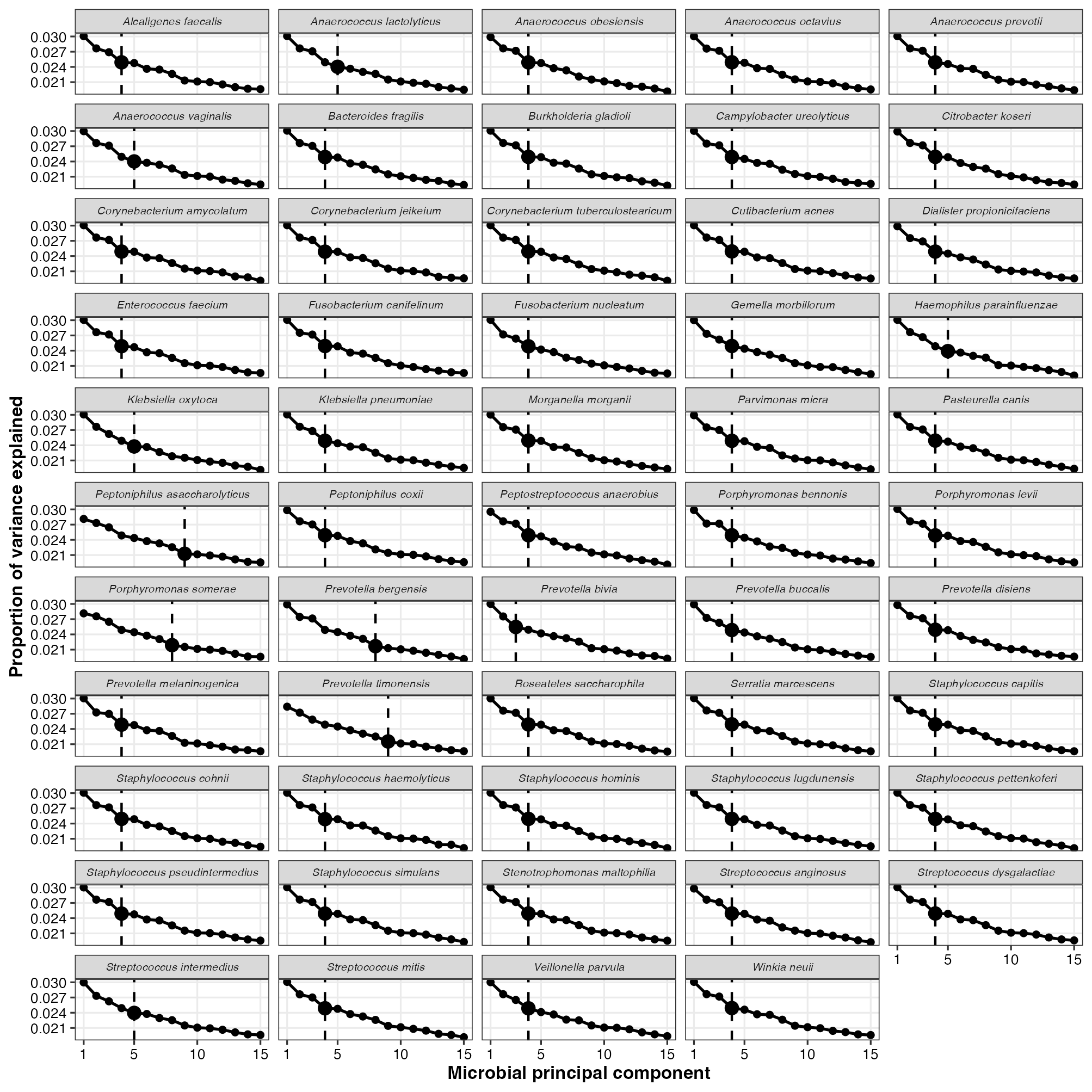

### Supplemental Figure 3

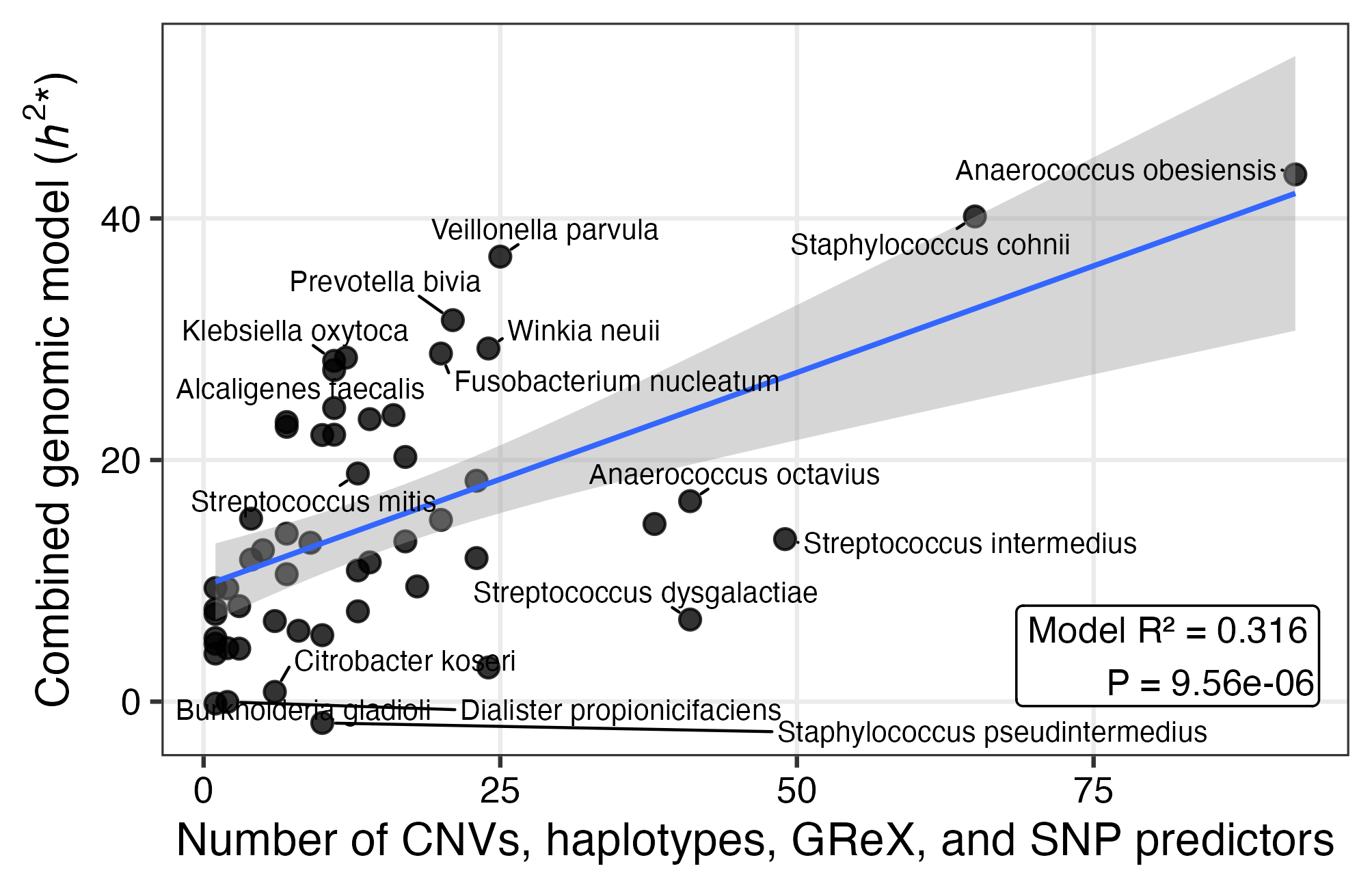
